# The mental wellbeing of prison staff in England during the COVID-19 pandemic: a cross-sectional study

**DOI:** 10.1101/2022.06.24.22276867

**Authors:** Luke Johnson, Maciej Czachorowski, Kerry Gutridge, Nuala McGrath, Julie Parkes, Emma Plugge

## Abstract

**Objectives:** To examine the mental wellbeing of prison staff in England within the pandemic, and determine factors associated with wellbeing.

**Design:** Cross-sectional study, with self-completed hardcopy and online surveys.

**Setting:** 26 prisons across England, chosen to be representative of the wider closed prison estate in England

**Participants:** All staff within the 26 prisons from 20^th^ July 2020 and 2^nd^ October 2020 were eligible.

**Primary outcome measure:** Wellbeing, measured using the Short-version of Warwick-Edinburgh Wellbeing Scale (SWEMWBS). Staff wellbeing was compared to that of the English population using indirectly standardised data from the Health Survey for England 2010-13 and a one-sample t-test. Multivariate linear regression modelling explored associations with mental wellbeing score.

**Results:** 2534 individuals were included (response rate 22.2%). The mean age was 44 years, 53% were female, and 93% were white. The sample mean SWEMWBS score was 23.84 and the standardised population mean score was 23.57. The difference in means was statistically significant (95% CI 0.09 to 0.46), but not of at a clinically meaningful level. The multivariate linear regression model was adjusted for age category, sex, ethnicity, smoking status, presence of comorbidities, occupation, and HMPPS region. Higher wellbeing was significantly associated with older age, male sex, Black/Black British ethnicity, never having smoked, working within the health staff team, and working in certain prison regions. The overall model had a low predictive value (adjusted R^2^ = 0.0345).

**Conclusions:** Unexpectedly, prison staff wellbeing as measured by SWEMWBS was similar to that of the general population. Reasons for this are unclear but could include the reduction in violence within prisons since the start of the pandemic. Qualitative research across a diverse sample of prison settings would enrich understanding of staff wellbeing within the pandemic.

**Strengths and limitations of this study:**

- This is the largest study to date to explore the mental wellbeing of prison staff in the UK (n=2534) and the first peer-reviewed study examining this during the COVID-19 pandemic.
- The sampling frame used (all staff at 26 prisons in England) is more likely to be representative of the prison staff population than other studies which have measured prison staff wellbeing and recruited through trade union channels
- Wellbeing was measured using SWEMWBS, which has been validated within the UK population
- Response rate was low (22.2%) and a number of variables adjusted for in the regression model were self-reported which could lead to a degree of bias

## Introduction

The prison system in England and Wales consists of 117 prisons, holding approximately 80,000 prisoners and employing approximately 53,000 staff.[1] Early in the COVID-19 pandemic, prisons were identified as high-risk environments for outbreaks because of overcrowded conditions and a high risk of severe COVID-19 in prison residents and staff.[2] In prisons across England, infection control measures implemented in March 2020 introduced a restricted regime for residents, with the cessation of social visits and activities, face-to-face education, and training and employment opportunities.[3] Restrictions on the numbers of people unlocked and numbers of people in exercise yards at any one time were also introduced to facilitate social distancing. However, these measures resulted in residents being confined to their cells for up to 23 hours a day.[2]

Prisons employ a diverse group of professionals, including prison officers, probation officers, administrators, nurses, doctors, psychologists, chaplains, and management. Most staff in English prisons are employed by Her Majesty’s Prison and Probation Service (HMPPS), though nurses and doctors are employed by the National Health Service (NHS), and some employees are contract workers (such as maintenance workers). Prior to the pandemic, the available evidence suggested that prison staff in England had low wellbeing and a high burden of mental health issues.[4-9] Factors associated with lower wellbeing included the demanding workload, high risk of violence and workplace injury, exposure to prison resident self-harm, and an oppressive work environment.[10, 11] Most prison staff are classed as essential workers and have continued working in prisons throughout the pandemic.[3] They have had to adapt to changes in prison regime, taking on new roles and responsibilities. By continuing to fulfil their duties at work, staff have put their own health, and that of their families, at risk.[2] Recent findings suggested a higher burden of mental health symptoms in UK frontline workers than the general population during the first month of the March 2020 lockdown.[12] Whilst it might be hypothesised that both the pandemic and changes to the prison regime have worsened wellbeing in prison staff in England,[2] there is little evidence regarding this impact.[2] The present study aims to examine the mental wellbeing of prison staff in England during the pandemic, and determine factors associated with wellbeing.

## Methods

This cross-sectional study uses data collected as part of the ‘COVID-19 in Prisons Study’ (CiPS). CiPS aimed to examine the epidemiology of SARS-CoV-2 in prisons in England to inform policy and practice during the pandemic and recovery period. One objective was to examine the mental wellbeing of staff.[13] CiPS consisted of a repeated panel survey in 28 prisons in England, selected for their representativeness of closed prisons in function, security category, geographical area, staff population, resident population, and prior COVID-19 outbreak.

Data from rounds 1 and 2 were collected between 20^th^ July 2020 and 2^nd^ October 2020. In each round, a questionnaire was administered to capture self-reported sociodemographics, height, weight, medical history, and symptoms (see supplementary material for questionnaire). Participants were also asked if they were on immune compromising medications and if a household member worked in a health or social care setting. The questionnaire collected the first four digits of a participant’s home postcode, which was used to estimate the index of multiple deprivation (IMD) based on the IMD category most prevalent in that postcode. Each prison provided information on the prison environment and the number of prison staff and residents. Staff wellbeing information was recorded in the second round, using the Short version of the Warwick Edinburgh Mental Wellbeing Scale (SWEMWBS), a wellbeing measure which has been validated in the UK population.[14, 15] All staff in the 28 prisons were eligible to participate if they met inclusion criteria (table 1).

**Table 1.** Inclusion and exclusion criteria for staff

| **Inclusion criteria** | **Exclusion criteria** |
| --- | --- |
| • Aged 18 years or older<br>• Currently employed in a study prison<br>• Willing and able to give informed, voluntary consent to study participation | • Unwilling or unable to give informed, voluntary consent for study participation<br>• Staff who are shielding or on long-term absence during the study period |

The range of possible scores for SWEMWBS is 7-35 and the minimum important level of difference in SWEMWBS score, defined as ‘of significance to the patient, member of the public, or the health professional’, has been estimated to be one point.[15] Using the sample SWEMWBS standard deviation (SD) (4·84), an α Of 0·05, and a power of 90%, it was determined post-hoc using nQuery 8·7·0·0 that sample size was sufficiently large to detect a difference of one point in SWEMWBS scores between groups in all performed statistical tests.[16]

### Patient and public involvement

HMPPS staff and NHS England and NHS Improvement (NHSEI) were involved in the development and conduct of the study. People with experience of imprisonment were also represented on the study steering group.

### Statistical analyses

Stata (version 16) was used for analysis.[17] Descriptive statistics were generated to characterise the sample. Characteristics (age and sex) of the participating sample were compared to the total staff population in prisons targeted for recruitment (the eligible population), as well as the total staff population in the English prison system. The sample mean SWEMWBS score was compared to the mean SWEMWBS score for the English population in the Health Survey for England (HSE) 2010-2013, using summary data from a previous academic publication which was first age-sex standardised to the participating staff population distribution.[14]

Univariable and multivariable linear regression models were used to identify associations with SWEMWBS score. Categorical variables were represented in models using dummy variables. Variables which were deemed clinically important a priori or showed statistical significance (p<0·05) at the univariable stage were considered for inclusion in the final model. Age, sex, ethnicity, smoking status, and presence of comorbidities were judged clinically important because of known associations with wellbeing.[14, 18] The multivariable linear regression model was created using backward selection, with all variables with one or more categories with a p<0·05 being included in the final model. For regression modelling, age was dichotomised into 40 years and under and 41 years and over to divide the population approximately in half. Model assumptions were checked, including the use of Cook’s distance to identify observations of high influence and leverage.

## Results

### Sample overview

Of 11409 staff eligible to participate, there were 2556 responses. Of these, 22 were removed because of missing answers to the SWEMWBS questionnaire, preventing calculation of the SWEMWBS score. A further two questionnaires which were completed but not linked to participant information were also excluded. 2534 responses are therefore included in this analysis (22·2% of eligible staff). Nearly all eligible prisons (26 of 28) had staff who participated in the SWEMWBS questionnaire. Of these 26 prisons, the proportion of eligible staff participating varied between 0·2% to 52·1%. Of staff participating in round two, 60·2% completed the SWEMWBS questionnaire (figure 1).

**Figure 1.**
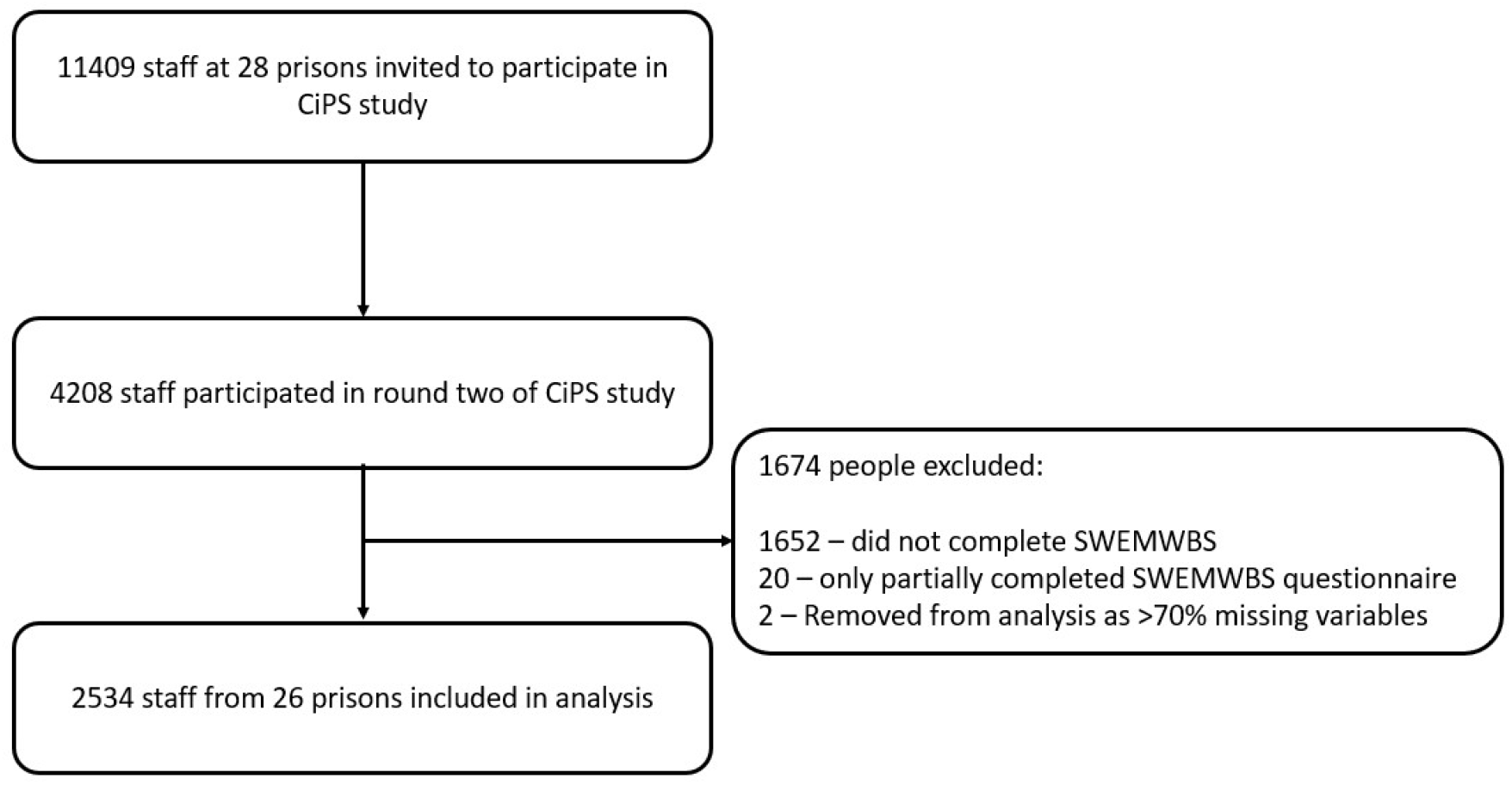
Survey flowchart: eligible population, recruitment, usable responses. Characteristics (age and sex) of the sample were compared with the total staff population in the 28 prisons (table 2), and the total staff population in the English prison estate (table 3). Females were overrepresented in the sample compared to each population (p<0·001 for both calculations), as were older staff members (p<0·001 for both calculations).

The mean age was 43·7 years (range 19 to 76; SD 12·1). Most participants were of white ethnicity (93.3%) and the majority lived in areas in the three least deprived quintiles (83·0%). A substantial minority (29·0%) lived with someone who worked in a health or social care setting. Just under half were either smokers or ex-smokers (44·9%). The majority (66·7%) self-reported they were overweight, obese or extremely obese, and 19·3% had one or more comorbidities.

The majority of staff directly worked for the prison service (79·3%) rather than a prison-related employer. Most participants worked in a male prison (89·0%); 8·9% worked in a female prison within the sample, and 2·1% worked in Young Offender Institutions (YOIs). Prisons were grouped by HMPPS region to take account of both geographical variation and variation in work culture – each region being managed by a Prison Group Director. Some regions only contained one prison in the sample, whilst others contained several. Unlike male prisons, female prisons and YOIs are not assigned security categories, and so are categorised separately within the prison security variable.

Descriptive statistics and mean SWEMWBS score in each category are presented in table 4. The mean SWEMWBS score was 23·84 (range 7 to 35; SD 4·84). The distribution of scores is shown in figure 2. The distribution was judged as normal but with four total SWEMWBS scores of elevated frequency. It was ascertained that these totals related to people who had marked all SWEMWBS questions with the same answer, either 1, 2, 3, or 5. A score of ≤17 is highly correlated with mental illness[19] – 4·4% (n=111) of participants scored ≤17.

**Table 2.**
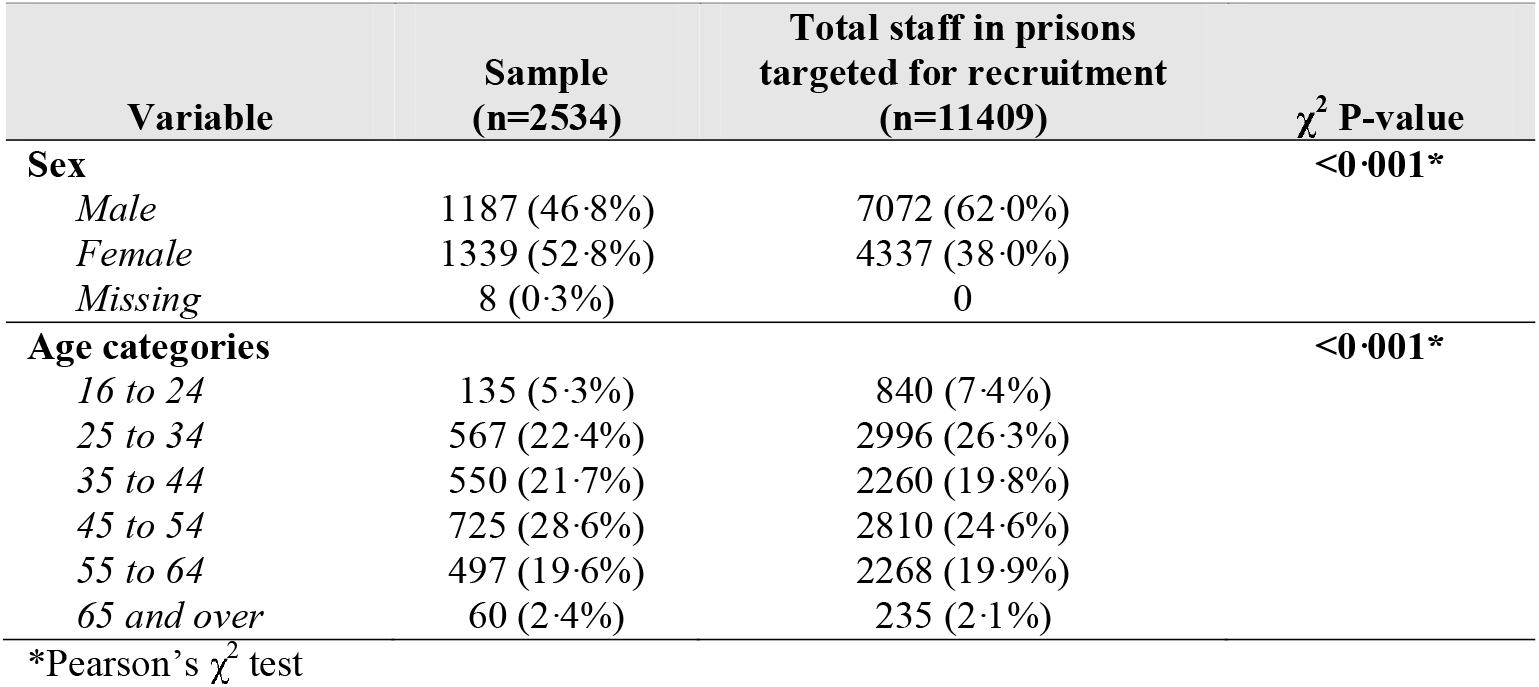
Comparison of age and sex distributions between sample and total staff in prisons targeted for recruitment

**Table 3.** Comparison of age and sex distributions between sample and total staff in English prison estate

| Variable | Sample (n=2534) | Total staff in prison estate (n = 53473) | $\chi^2$ P-value |
| --- | --- | --- | --- |
| **Sex** |  |  | **<0.001\*** |
| Male | 1187 (46.8%) | 27049 (50.6%) |  |
| Female | 1339 (52.8%) | 26424 (49.4%) |  |
| Missing | 8 (0.3%) | 0 |  |
| **Age categories** |  |  | **<0.001\*** |
| 16 to 24 | 135 (5.3%) | 3456 (6.5%) |  |
| 25 to 34 | 567 (22.4%) | 13565 (25.4%) |  |
| 35 to 44 | 550 (21.7%) | 11926 (22.3%) |  |
| 45 to 54 | 725 (28.6%) | 13036 (24.4%) |  |
| 55 to 64 | 497 (19.6%) | 10427 (19.5%) |  |
| 65 and over | 60 (2.4%) | 1063 (2.0%) |  |

\*Pearson's $\chi^2$ test

**Table 4.** Characteristics of sample and SWEMWBS score

| Variable (n=2534) | Distribution<br>N (%) | SWEMWBS Score<br>Mean (SD) |
| --- | --- | --- |
| **Age category (years)** |  |  |
| ≤30 | 464 (18.3%) | 23.33 (4.61) |
| 31-40 | 580 (22.9%) | 23.50 (4.74) |
| 41-50 | 613 (24.2%) | 23.79 (4.89) |
| 51-60 | 706 (27.8%) | 24.31 (4.95) |
| ≥61 | 174 (6.8%) | 24.64 (4.90) |
| Missing | 0 |  |
| **Sex** |  |  |
| Male | 1187 (46.8%) | 24.32 (5.14) |
| Female | 1339 (52.8%) | 23.43 (4.53) |
| Missing | 8 (0.3%) |  |
| **Ethnicity** |  |  |
| White | 2364 (93.3%) | 23.78 (4.80) |
| Black/Black British | 68 (2.7%) | 25.27 (5.26) |
| Asian/Asian British | 48 (1.9%) | 24.66 (5.65) |
| Other ethnicity | 53 (2.1%) | 24.13 (4.80) |
| Missing | 1 (0.0%) |  |
| **BMI Category** |  |  |
| Underweight (<18.5 kg/m<sup>2</sup>) | 18 (0.7%) | 24.54 (5.16) |
| Healthy (18.5-24.9 kg/m<sup>2</sup>) | 658 (26.0%) | 23.72 (4.49) |
| Overweight (25-29.9 kg/m<sup>2</sup>) | 941 (37.1%) | 23.74 (4.84) |
| Obese (30-39.9 kg/m<sup>2</sup>) | 672 (26.5%) | 24.17 (5.01) |
| Extremely obese (≥40 kg/m<sup>2</sup>) | 78 (3.1%) | 23.65 (4.87) |
| *Missing* | 167 (6.6%) |  |
| **Smoking status** |  |  |
| *Current smoker* | 344 (13.6%) | 23.46 (5.08) |
| *Ex-smoker* | 785 (31.0%) | 23.64 (4.97) |
| *Never smoked* | 1396 (55.1%) | 24.07 (4.70) |
| *Missing* | 9 (0.4%) |  |
| **Number of comorbidities** |  |  |
| *0* | 2046 (80.7%) | 23.90 (4.83) |
| *1 or more* | 488 (19.3%) | 23.61 (4.87) |
| **Immune compromising medications** |  |  |
| *Yes* | 113 (4.5%) | 23.96 (5.01) |
| *No* | 2390 (94.3%) | 23.84 (4.83) |
| *Missing* | 31 (1.2%) |  |
| **Occupation** |  |  |
| *Prison service staff* | 2009 (79.3%) | 23.75 (4.90) |
| *Health staff* | 197 (7.8%) | 24.51 (4.89) |
| *Agency staff* | 259 (10.2%) | 24.16 (4.69) |
| *Probation service* | 66 (2.6%) | 23.33 (2.99) |
| *Missing* | 3 (0.1%) |  |
| **IMD Quintile of staff residence** |  |  |
| *1<sup>st</sup> and 2<sup>nd</sup> quintiles (most deprived)* | 425 (16.8%) | 23.86 (4.88) |
| *3<sup>rd</sup> quintile* | 1086 (42.9%) | 23.74 (4.77) |
| *4<sup>th</sup> quintile* | 870 (34.3%) | 24.07 (4.87) |
| *5<sup>th</sup> quintile (least deprived)* | 148 (5.8%) | 23.21 (5.11) |
| *Missing* | 5 (0.2%) |  |
| **Household member working in a health or social care setting** |  |  |
| *Yes* | 735 (29.0%) | 23.79 (4.95) |
| *No* | 1791 (70.7%) | 23.86 (4.79) |
| *Missing* | 8 (0.3%) |  |
| **HMPPS Region** |  |  |
| *R1* | 176 (7.0%) | 23.18 (4.93) |
| *R2* | 292 (18.5%) | 23.95 (4.51) |
| *R3* | 594 (23.4%) | 23.94 (5.06) |
| *R4* | 486 (19.2%) | 23.34 (4.10) |
| *R5* | 69 (2.7%) | 26.59 (5.67) |
| *R6* | 89 (3.5%) | 23.61 (4.48) |
| *R7* | 67 (2.6%) | 23.01 (4.54) |
| *R8* | 1 (0.0%) | 25.03 (n/a) |
| *R9* | 52 (2.1%) | 22.31 (4.25) |
| *R10* | 99 (3.9%) | 24.11 (4.71) |
| *R11* | 176 (7.0%) | 24.92 (5.46) |
| *R12* | 175 (6.9%) | 23.65 (4.76) |
| *R13* | 258 (10.2%) | 24.07 (5.24) |
| *Missing* | 0 |  |
| **Prison security category** |  |  |
| *A* | 709 (28.0%) | 24.06 (5.17) |
| *B* | 769 (30.4%) | 23.83 (4.73) |
| *C* | 778 (30.7%) | 23.61 (4.56) |
| *Female prisons* | 225 (8.9%) | 24.34 (5.10) |
| *YOIs* | 53 (2.1%) | 22.36 (4.23) |
| *Missing* | 0 |  |
| **Prison functional category** |  |  |
| *Trainer* | 1624 (64.1%) | 23.91 (4.92) |
| *Local* | 858 (33.9%) | 23.81 (4.70) |
| *YOIs* | 53 (2.1%) | 22.36 (4.23) |
| *Missing* | 0 |  |
| **Prisoner/staff ratio** |  |  |
| *≤ 1* | 643 (25.4%) | 23.51 (4.92) |
| *1-2* | 1016 (40.1%) | 24.28 (4.95) |
| $\geq 2$ | 875 (34.5%) | 23.58 (4.60) |
| Missing | 0 |  |

Abbreviations: BMI: Body Mass Index; IMD: Index of Multiple Deprivation; HMPPS: Her Majesty's Prison and Probation Service; SD: Standard deviation; YOI: Young Offender Institutions

**Figure 2.**
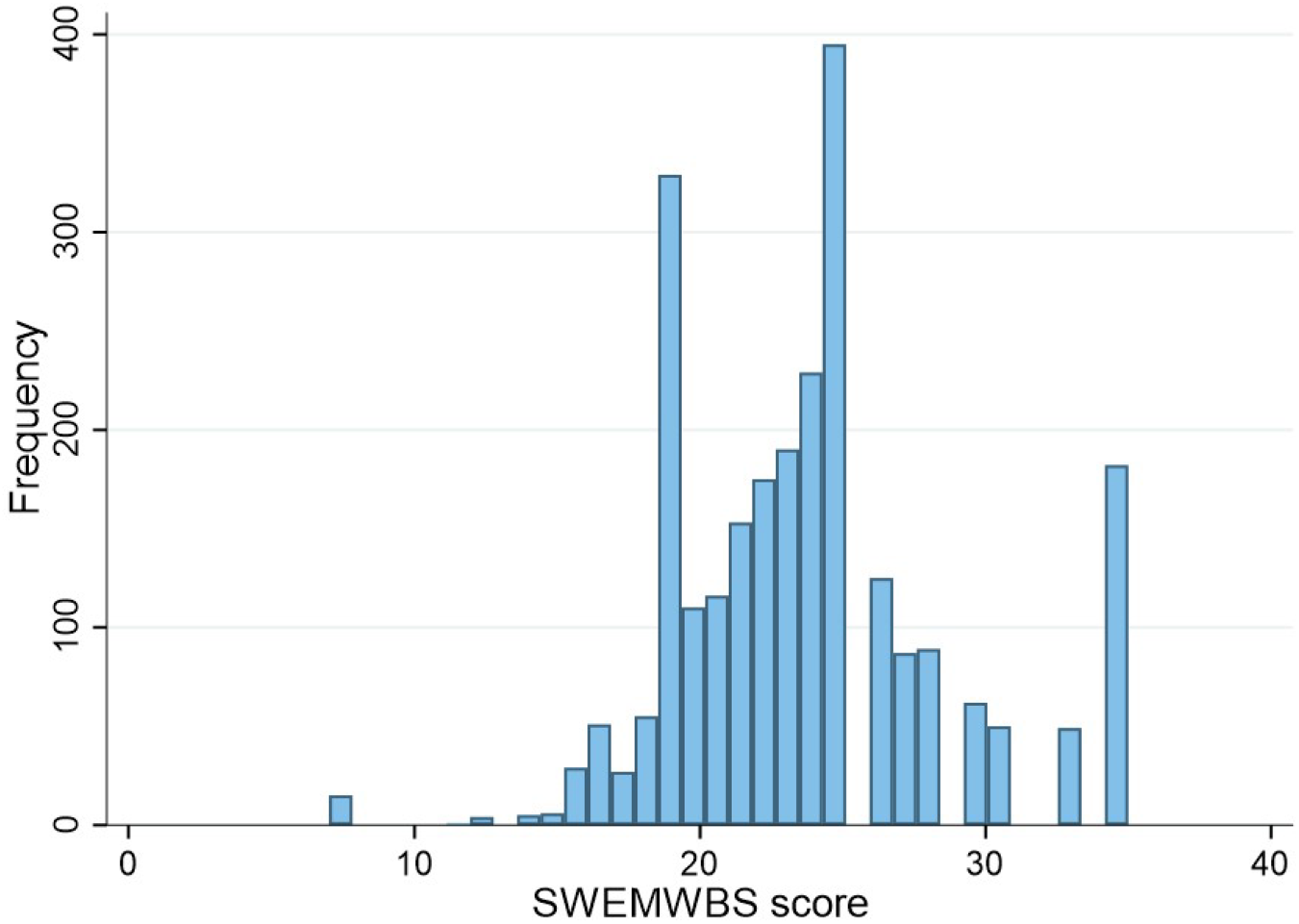
Histogram of SWEMWBS variable. The HSE standardised mean SWEMWBS score was 23·57. The prison staff sample had a statistically significantly higher mean SWEMWBS score (23·84) than the standardised HSE mean (p=0·004; 95% CI 23·66, 24·03). However, this was less than the previously defined important difference of one point.

In univariate linear regression analysis, SWEMWBS score was significantly associated with age, sex, ethnicity, smoking status, occupation, HMPPS region, prisoner functional category, and prisoner/staff ratio. The final multivariable linear regression model was adjusted for age category, sex, ethnicity, smoking status, presence of comorbidities, occupation, and HMPPS region (table 5). Significantly higher SWEMWBS scores were associated with older age, male sex, Black/Black British ethnicity and staff occupation (specifically health staff compared to prison service staff). Additionally, HMPPS regions R5 and R11 were significantly associated with higher wellbeing as compared to R1 at the univariable and multivariable level (at multivariate level, β 3·81 (95% CI 2·47, 5·15, p<0·001) and β 1·93 (95% CI 0·93, 2·94, p<0·001), respectively). The only definitive meaningfully important difference in score (where confidence intervals did not cross minus one or one) came from working in the R5 as compared to R1 (95% CI 2·47, 5·15). The multivariate model did not explain the majority of variance (R^2^ = 0·0429).

**Table 5.** Primary model: Unadjusted and adjusted linear regression models for predicting SWEMWBS score

|  | Unadjusted models (N=2512) |  |  | Adjusted model<sup>I</sup> (N=2512) |  |  |
| --- | --- | --- | --- | --- | --- | --- |
| | $\beta^*$ | 95% CI | P-value | $\beta^*$ | 95% CI | P-value |
| **Age category** |  |  |  |  |  |  |
| 40 and under | 0 |  |  | 0 |  |  |
| 41 and over | 0.70 | (0.32, 1.09) | **<0.001** | 0.53 | (0.14, 0.92) | **0.008** |
| **Sex** |  |  |  |  |  |  |
| Female | 0 |  |  | 0 |  |  |
| Male | 0.90 | (0.52, 1.28) | **<0.001** | 0.95 | (0.53, 1.37) | **<0.001** |
| **Ethnicity** |  |  |  |  |  |  |
| White | 0 |  |  | 0 |  |  |
| Black/Black British | 1.62 | (0.42, 2.83) | **0.008** | 1.45 | (0.25, 2.66) | **0.018** |
| Asian/Asian British | 0.87 | (-0.51, 2.26) | 0.218 | 0.63 | (-0.74, 2.00) | 0.369 |
| Other ethnicity | 0.34 | (-0.98, 1.66) | 0.610 | 0.30 | (-1.01, 1.60) | 0.656 |
| **Smoking status** |  |  |  |  |  |  |
| *Never smoked* | 0 |  |  | 0 |  |  |
| *Current smoker* | -0.62 | (-1.19, -0.04) | **0.036** | -0.51 | (-1.08, 0.07) | 0.083 |
| *Ex-smoker* | -0.44 | (-0.87, -0.02) | **0.041** | -0.44 | (-0.87, -0.02) | **0.040** |
| **No. of comorbidities** |  |  |  |  |  |  |
| *None* | 0 |  |  | 0 |  |  |
| *1 or more* | -0.31 | (-0.79, 0.18) | 0.213 | -0.41 | (-0.89, 0.07) | 0.091 |
| **Occupation** |  |  |  |  |  |  |
| *Prison service staff* | 0 |  |  | 0 |  |  |
| *Health staff* | 0.77 | (0.05, 1.48) | **0.035** | 1.32 | (0.60, 2.05) | **<0.001** |
| *Agency staff* | 0.42 | (-0.21, 1.05) | 0.192 | 0.62 | (-0.01, 1.25) | 0.053 |
| *Probation staff* | -0.43 | (-1.62, 0.76) | 0.479 | 0.06 | (-1.13, 1.24) | 0.923 |
| **HMPPS Region** |  |  |  |  |  |  |
| *R1* | 0 |  |  | 0 |  |  |
| *R2* | 0.82 | (-0.08, 1.73) | 0.074 | 0.86 | (-0.04, 1.76) | 0.062 |
| *R3* | 0.80 | (-0.01, 1.61) | 0.054 | 0.87 | (0.05, 1.69) | **0.038** |
| *R4* | 0.19 | (-0.64, 1.02) | 0.656 | 0.31 | (-0.52, 1.14) | 0.465 |
| *R5* | 3.43 | (2.09, 4.78) | **<0.001** | 3.81 | (2.47, 5.15) | **<0.001** |
| *R6* | 0.46 | (-0.77, 1.69) | 0.465 | 0.67 | (-0.55, 1.89) | 0.280 |
| *R7* | -0.15 | (-1.50, 1.21) | 0.832 | 0.06 | (-1.30, 1.41) | 0.932 |
| *R9* | -0.76 | (-2.27, 0.74) | 0.319 | -0.60 | (-2.09, 0.89) | 0.431 |
| *R10* | 0.96 | (-0.23, 2.14) | 0.115 | 0.91 | (-0.27, 2.09) | 0.130 |
| *R11* | 1.77 | (0.76, 2.78) | **0.001** | 1.93 | (0.93, 2.94) | **<0.001** |
| *R12* | 0.52 | (-0.49, 1.53) | 0.316 | 0.57 | (-0.43, 1.58) | 0.263 |
| *R13* | 0.92 | (-0.01, 1.85) | 0.051 | 1.01 | (0.08, 1.94) | **0.032** |
| **Prisoner functional category** |  |  |  |  |  |  |
| *Trainer* | 0 |  |  | - |  |  |
| *Local* | -0.11 | (-0.51, 0.30) | 0.606 |  |  |  |
| *YOIs* | -1.53 | (-2.88, -0.18) | **0.018** |  |  |  |
| **Prisoner/staff ratio** |  |  |  |  |  |  |
| *≤ 1* | 0 |  |  | - |  |  |
| *1-2* | 0.79 | (0.31, 1.26) | **0.001** |  |  |  |
| *> 2* | 0.08 | (-0.42, 0.57) | 0.761 |  |  |  |

<sup>1</sup>Adjusted for age category, sex, ethnicity, smoking status, number of comorbidities, occupation, and HMPPS region.
\*Beta coefficient for dependent outcome: WEMWBS Score.
Abbreviations: CI: Confidence interval; YOI: Young Offender Institution

When checking model assumptions,159 observations were deemed influential using Cook’s distance values (values greater than 4/n, where n is the number of observations (>0·00159) were considered influential). Of these, 114 (71·7%) were SWEMWBS scores of 7 or 35 – people who had scored one or five for all questions. Another model (see table six in supplementary material) was therefore created excluding observations with a SWEMWBS score of 7 or 35. The same covariates were included to enable comparability with the primary model. Removing the influential observations caused resulted in no important changes to the estimates in table 5.

## Discussion

This is one of the largest studies of prison staff health in England and it showed that the wellbeing of prison staff in England during the COVID-19 pandemic was similar to that of the general population pre-pandemic. Prison staff wellbeing was statistically significantly higher than that of the general population but this was not considered clinically meaningful, using the threshold of one-point on the SWEMWBS scale. It is important to note that we used data from the HSE 2010-13[14] as a comparison and it is likely that wellbeing among the general population has since changed because of the pandemic; SWEMWBS data from an online, quota-based questionnaire collected from March to May 2020 – at the start of the first lockdown – suggested that general population wellbeing within the UK was lower than that at the time of the HSE 2010-13.[20] Therefore, the difference in wellbeing may be larger in comparison to the general population.

Within the prison staff population, higher wellbeing was associated with older age, male sex, Black/Black British ethnicity, never having smoked, working within the health staff team, and working in certain prison regions. Working in the R5 region represented the only meaningfully important improvement in wellbeing. Differences in wellbeing between HMPPS regions could be the consequence of geographical variation or, more likely, differences in aspects of workplace culture identified as important to prison staff wellbeing in previous systematic reviews.[10, 11] Key aspects identified within these reviews include support from management, workload, and clarity of job role.

This study is the largest to date to explore the wellbeing of prison staff in the UK and one of only two examining their wellbeing during the COVID-19 pandemic.[21] It uses SWEMWBS, a measure of wellbeing that has been validated in the UK population.[14, 22] The sample was well-powered to detect meaningfully important differences in wellbeing. As far as the authors are aware, it is the first quantitative study in the UK to measure prison staff wellbeing across multiple prisons without using trade union membership as a sampling frame, which may bias the results as they represent only one sector of staff

The low response rate of this study (22·2%) may have caused some selection bias; participants were more likely to be female and older than the population of all prison staff, and female and older prison staff were found to have higher levels of wellbeing. Two prisons did not provide any responses to the SWEMWBS questionnaire because of a miscommunication. Additionally, certain SWEMWBS scores were overly common within the expected distribution. These represented participants who had scored all questionnaire answers the same. This has not been reported in other studies using SWEMWBS, and it is uncertain why this happened here. However, a sensitivity analysis removing those who answered all questions as the lowest or highest value did not substantially alter results. Further, the study did not gather information on pre-existing mental illness, which might have helped improve the fit of the regression model, as mental health has been shown to be associated with SWEMWBS score.[20]

This study found that prison staff wellbeing during the pandemic was similar to that of the general population prior to the pandemic. Only 4·4% of staff scores indicated possible mental illness. This contrasts with other cross-sectional studies from prior to the pandemic which, using the General Health Questionnaire, estimated 56·6-95% of UK prison officers met criteria for potential mental illness.[4-7] Two of these studies were conducted in a single prison setting, whereas the other two used an online survey conducted through trade union channels. Findings also deviate from another study of UK prison officer wellbeing during the pandemic, in which 43% had symptoms of moderate or severe anxiety.[21] However, the latter study suffered significant risk of bias because recruitment was conducted by email, through trade union channels, with a response rate of only 2·0%. Furthermore, with the exception of the GAD-7, none of the study measures were validated, and the study itself was not peer-reviewed.

Reasons for the variations in results between this study and studies prior to the pandemic are unclear and could include the aforementioned differences in study methodology or a true difference in prison staff wellbeing. Such a difference in prison staff wellbeing could have been brought about through pandemic-related changes to prison job roles and operations. Prison residents have spent large periods within their cells over the past 18 months, which has adversely impacted their mental health.[2] However, it is also likely to have greatly impacted the role of prison staff.[3] Moreover, the prison population in England and Wales has reduced by more than 5% since the start of the pandemic, with possible consequences on staff workload.[1] Job demands and role clarity have been shown to be two key factors influencing staff wellbeing.[5, 7, 23] Although the role of staff has likely been altered multiple times throughout the pandemic, the decrease in prison population and restriction in prison resident movement and activities may have led to a reduction in staff duties. This is likely to have contributed to the decline in assaults within prisons in England and Wales, which decreased by 40% in the 12 months following March 2020.[24] The number of serious assaults decreased by 47% and assaults on staff decreased by 24%. These reductions will likely have had a positive effect on staff wellbeing.[23] Numbers of self-harm incidents recorded in England and Wales have reduced 22% in male prisons and 4% in female prisons within the same period. This change is partially the result of the reduction in prison populations – rates of self-harm incidents have only reduced 19% in male residents and, in fact, have increased 12% in female residents. Nevertheless, fewer self-harm incidents may also have benefited staff wellbeing.[25-27]

Similar to the HSE 2010-13, within the multivariate model, SWEMWBS score was significantly higher for people of black ethnicity, older age, and non-smokers.[14] Many of the factors previously found to be strongly associated with prison staff wellbeing were not measured directly within this study, perhaps explaining the model’s lack of predictive ability. Such factors included job demands,[5, 7] support from management,[6] relationships with colleagues,[5, 6], role clarity,[5] detachment,[28] and experiences of aggression.[28] Some of these may be mediating the differences found between health staff and other prison staff, and between different prison regions. Risk factors for COVID-19 mortality were not associated with worse wellbeing. Index of multiple deprivation and BMI were not significantly associated with wellbeing, and older age and male sex were associated with increased wellbeing.

As prisons resume normal functioning, it is important that staff wellbeing is protected and improved. In particular, job role and demands should be considered, and whole-system efforts to address self-harm and assault should be reinvigorated to minimise the adverse impacts on both prison residents and staff. Focus should be placed on continuing to reduce the imprisoned population to ensure the safety of prison staff and residents and enabling the development of a genuinely rehabilitative culture. This is particularly important considering the lack of evidence that imprisonment reduces rates of recidivism,[29] and that most people are imprisoned for non-violent offences, so are unlikely to pose a risk to the public.[30].

Further research is needed to build on the findings of this study. Qualitative research across a broad range of prison staff groups and occupations would enrich what is currently known about prison staff wellbeing and the impact of the pandemic in prisons. Such research could look at the experiences of staff whilst working in the pandemic, and any changes in perception of their role or relationship to prison residents as a result.

Additionally, more evidence is needed on interventions to improve prison staff wellbeing. In collaboration with key organisations such as HMPPS and the Prison Officers Associations, research across these areas would help to build more successful strategies for promoting staff wellbeing and lead to a culture of shared learning.

## Data Availability

The dataset used in this study is held securely on encrypted servers at the University of Southampton and all data is managed in accordance with the CiPS Privacy Notice and study protocol, as well as the General Data Protection Regulation (GDPR) guidelines. The safeguarding of participant confidentiality is of utmost importance and therefore any requests for data will be judged on a case-by-case basis by all relevant stakeholders. All shared data would be deidentified. For all requests regarding data, please contact the corresponding author.

## Ethics statement

Ethical approval was granted by the University of Southampton (ERGO ID 57844) and the Health Research Authority and Health and Care Research Wales (IRAS project ID: 285534; REC reference: 20/NW/0320).

## Acknowledgements

We would like to thank colleagues at the Department of Health and Social Care, Office for National Statistics, Ministry of Justice, Her Majesty’s Prison and Probation Service, UK Health Security Agency, and NHS England and NHS Improvement for their assistance.

## Contributions

JP, NM, NC and EP designed the study. JP, NM, NC, EP and KG were involved in study implementation. Analysis was undertaken by LJ, MC and NM. LJ and EP drafted the manuscript, and all authors were involved in revising it. All authors approved the final manuscript.

The corresponding author (LJ) affirms that the manuscript is an honest, accurate, and transparent account of the study being reported; that no important aspects of the study have been omitted; and that any discrepancies from the study as planned have been explained.

