## Supplementary material for "The mental wellbeing of prison staff in England during the COVID-19 pandemic: a cross-sectional study"

**Contents**

Page 2 – Study questionnaire for staff

Page 5 – Secondary model for predicting SWEMWBS scores

### Study Questionnaire for Staff

Public Health England

Protecting and improving the nation's health

**Short Study Title: COVID-19 in Prisons Study (CiPS)**

**Staff Questionnaire**

Thank you for agreeing to fill in this questionnaire.

Please try to answer every question even though some may seem rather similar to others, or may not seem relevant to you.

This is not a test. There are no "right" answers. Please tick the box that best describes your own experience.

The information you give on this questionnaire is anonymous and will be treated in the strictest confidence.

**Thank you for your help with this study.**

QUESTIONS

1. What is the URN on your swab test?
Please enter the URN again

2. What is your date of birth?
3. What is your sex?
4. What is your ethnic group?
(Choose one option that best describes your ethnic group or background)

White
- English / Welsh / Scottish / Northern Irish / British
- Irish
- Gypsy or Irish Traveller
- Any other White background, please describe

Mixed / Multiple ethnic groups
- White and Black Caribbean
- White and Black African
- White and Asian
- Any other Mixed / Multiple ethnic background, please describe

Asian / Asian British
- Indian
- Pakistani
- Bangladeshi
- Chinese
- Any other Asian background, please describe

Black / African / Caribbean / Black British
- African
- Caribbean
- Any other Black / African / Caribbean background, please describe

Other ethnic group

Arab

Any other ethnic group, please describe

5. What best describes your current grade/position within the prison? Please tick one box

- ☐ Prison Service Staff (Prison Officer)
- ☐ Prison Service Staff (Other)
- ☐ GFSL Contractors
- ☐ GFSL Cleaners
- ☐ NHS Staff
- ☐ Probation Service
- ☐ Psychological Services
- ☐ Other Agency Worker

6. What are the ***first*** 4 letters/digits of your home postcode?

7. Do you live in a household where there is also a person who is working or has worked during 2020, in a health or social care setting? (e.g. a nurse, carer, other prison officer etc)?
If yes, please state which occupation(s) Yes No

8. Do you have any of the following symptoms **today**? Please tick all that apply.

- ☐ Fever
- ☐ Muscle ache
- ☐ Fatigue
- ☐ Sore throat
- ☐ Cough
- ☐ Shortness of breath
- ☐ Headache
- ☐ Nausea and/or vomiting
- ☐ Abdominal pain
- ☐ Diarrhoea
- ☐ Loss of taste
- ☐ Loss of smell

9. Do you think/know you have **ever** been infected by SARS-CoV-2?

Yes No

**If yes**, what date did your symptoms start?

**If yes,** did you experience any of the following symptoms? (tick all that apply)

- ☐ Fever?
- ☐ Muscle ache?
- ☐ Fatigue?
- ☐ Sore throat?
- ☐ Cough?
- ☐ Shortness of breath?
- ☐ Headache?
- ☐ Nausea and/or vomiting?
- ☐ Abdominal pain?
- ☐ Diarrhoea?
- ☐ Loss of taste?

- ☐ Loss of smell?

**If yes**, were you tested? Yes No
**If yes**, were you positive? Yes No

10. Have you **ever** been tested for COVID-19?
    **If, yes**, please give date
    **If yes,** please state through which route
    A testing kit was sent to my home
    I attended a drive-in centre
    I was part of a research study
    Other, please state

11. Do you have any of the following health issues? Please tick all that apply.
    - ☐ Asthma
    - ☐ Chronic obstructive pulmonary disease (COPD), bronchitis or emphysema
    - ☐ Diabetes
    - ☐ Chronic kidney disease
    - ☐ Chronic liver disease
    - ☐ A neurological disease, for example, Parkinson's disease, multiple sclerosis
    - ☐ Heart disease
    - ☐ An organ transplant
    - ☐ Treatment for cancer (at the moment)
    - ☐ Bone marrow or stem cell transplant in the past 6 months
    - ☐ Any condition that means you have a very high risk of getting infections, such as SCID, sickle cell

12. Are you taking medicine that makes you more likely to get infections (such steroids)? Yes No

13. Do you smoke tobacco?
    Yes, current smoker
    No, ex-smoker
    No, never smoked

14. What is your height?

15. What is your weight?

THANK YOU FOR COMPLETING THE QUESTIONNAIRE

### Secondary model for predicting SWEMWBS scores

**Table 6. Model two: Unadjusted and adjusted linear regression models for predicting SWEMWBS score (excluding SWEMWBS scores of 7 and 35)**

| | Unadjusted models | | | Adjusted model[1] (N=2315) | | |
|---|---|---|---|---|---|---|
| | β* | 95% CI | P-value | β* | 95% CI | P-value |
| **Age category** | | | | | | |
| *40 and under* | 0 | | | 0 | | |
| *41 and over* | 0·61 | (0·31, 0·91) | **<0·001** | 0·52 | (0·21, 0·83) | **0·001** |
| **Sex** | | | | | | |
| *Female* | 0 | | | 0 | | |
| *Male* | 0·62 | (0·32, 0·91) | **<0·001** | 0·63 | (0·33, 0·94) | **<0·001** |
| **Ethnicity** | | | | | | |
| *White* | 0 | | | 0 | | |
| *Black/Black British* | 1·30 | (0·32, 2·27) | **0·013** | 1·07 | (0·09, 2·05) | **0·032** |
| *Asian/Asian British* | -0·16 | (-1·29, 1·22) | 0·784 | -0·32 | (-1·44, 0·80) | 0·773 |
| *Other ethnicity* | 0·19 | (-0·84, 1·22) | 0·719 | 0·15 | (-0·87, 1·17) | 0·580 |
| **Smoking status** | | | | | | |
| *Never smoked* | 0 | | | 0 | | |
| *Current smoker* | -0·65 | (-1·10, -0·20) | **0·005** | -0·63 | (-1·08, -0·18) | **0·006** |
| *Ex-smoker* | -0·27 | (-0·60, 0·07) | 0·118 | -0·31 | (-0·64, 0·02) | 0·065 |
| **No. of comorbidities** | | | | | | |
| *None* | 0 | | | 0 | | |
| *1 or more* | -0·28 | (-0·66, 0·10) | 0·149 | -0·37 | (-0·75, 0·00) | 0·052 |
| **Occupation** | | | | | | |
| *Prison service staff* | 0 | | | 0 | | |
| *Health staff* | 0·77 | (0·21, 1·33) | **0·007** | 1·08 | (0·51, 1·65) | **<0·001** |
| *Agency staff* | 0·40 | (-0·09, 0·90) | 0·109 | 0·49 | (0·00, 0·99) | 0·051 |
| *Probation staff* | 0·17 | (-0·73, 1·07) | 0·708 | 0·53 | (-0·38, 1·43) | 0·254 |
| **HMPPS Region** | | | | | | |
| *R1* | 0 | | | 0 | | |
| *R2* | 1·04 | (0·34, 1·75) | **0·004** | 1·07 | (0·37, 1·77) | **0·003** |
| *R3* | 0·86 | (0·22, 1·50) | **0·008** | 0·98 | (0·33, 1·62) | **0·003** |
| *R4* | 0·63 | (-0·02, 1·28) | 0·058 | 0·75 | (0·11, 1·40) | **0·023** |
| *R5* | 2·07 | (0·96, 3·18) | **<0·001** | 2·33 | (1·23, 3·44) | **<0·001** |
| *R6* | 0·41 | (-0·55, 1·37) | 0·401 | 0·56 | (-0·39, 1·52) | 0·249 |
| *R7* | 0·12 | (-0·95, 1·18) | 0·832 | 0·35 | (-0·72, 1·41) | 0·522 |
| *R9* | -0·77 | (-1·94, 0·40) | 0·196 | -0·60 | (-1·77, 0·56) | 0·310 |
| *R10* | 0·91 | (-0·03, 1·84) | 0·057 | 0·97 | (0·04, 1·90) | **0·040** |
| *R11* | 0·64 | (-0·17, 1·45) | 0·121 | 0·76 | (-0·04, 1·57) | 0·064 |
| *R12* | 0·38 | (-0·41, 1·17) | 0·348 | 0·41 | (-0·38, 1·19) | 0·312 |
| *R13* | 1·07 | (0·34, 1·80) | **0·004** | 1·22 | (0·49, 1·95) | **0·001** |
| **Prisoner functional category** | | | | | | |
| *Trainer* | 0 | | | - | | |
| *Local* | -0·03 | (-0·34, 0·29) | 0·861 | | | |
| *YOIs* | -1·52 | (-2·57, -0·48) | **0·004** | | | |
| **Prisoner/staff ratio** | | | | | | |
| *≤ 1* | 0 | | | - | | |
| *1-2* | 0·47 | (0·09, 0·85) | **0·014** | | | |
| *> 2* | 0·27 | (-0·12, 0·66) | 0·176 | | | |

[1]Adjusted for age category, sex, ethnicity, smoking status, number of comorbidities, occupation, and HMPPS region.

[*]Beta coefficient for dependent outcome: WEMWBS Score.

Abbreviations: CI: Confidence interval; YOI: Young Offender Institution
